# São Tomé and Príncipe on the verge of eliminating lymphatic filariasis as a public health problem: evidence from IDA impact assessment surveys

**DOI:** 10.64898/2026.06.15.26355657

**Authors:** Didier Bakajika, Alzíra Rosário, Jessica Vicente Fernandes, Lamine Diawara, Jean-Paul Tambwe Managala, Jean Kalenga, Vilfrido Santana Gil, Dyesse Yumba, Honorat M. Zouré, Elizabeth Juma, Abdoulaye Diarra, Jonathan King, Jorge Cano

## Abstract

**Background:** Accelerated efforts to eliminate lymphatic filariasis (LF) as a public health problem have been supported by the introduction of the triple-drug regimen of ivermectin, diethylcarbamazine and albendazole (IDA) in endemic settings. In São Tomé and Príncipe, nationwide mass drug administration (MDA) with diethylcarbamazine and albendazole was implemented in 2018, followed by IDA in 2019 and 2020. This study assesses progress towards elimination using post-MDA impact assessment surveys conducted after cessation of treatment.

**Methods:** Cross-sectional surveys were conducted among adults aged 20 years and older in 2022 and again between December 2024 and January 2025. Circulating filarial antigen (CFA) was detected using the filarial test strip (FTS). Individuals who tested positive were examined for microfilaremia using nocturnal calibrated thick blood smear microscopy. Additionally, programme data on MDA coverage and morbidity were obtained from national surveillance records.

**Results:** Three rounds of nationwide MDA achieved high epidemiological coverage (86.4% in 2018, 74.2% in 2019 and 80.0% in 2020). The impact assessment surveys conducted in 2022 evaluated 14 132 adults, with 21 individuals (0.15%) testing positive for CFA, while the follow-up survey conducted between December 2024 and January 2025 assessed 14 653 adults and detected seven positive cases (0.05%). No microfilariae were detected among the 28 antigen-positive individuals examined using nocturnal calibrated thick blood smears. National morbidity records documented 190 cases of lymphoedema and nine cases of hydrocoele.

**Conclusions:** Infection indicators remain well below WHO decision thresholds, suggesting that LF transmission is unlikely to be sustained. São Tomé and Príncipe appears to be close to eliminating LF as a public health problem. However, strengthening morbidity management services will be essential to support the preparation of the national elimination dossier.

**Author summary:** Lymphatic filariasis is a parasitic disease transmitted by mosquitoes that can cause chronic swelling of the limbs (lymphoedema) and scrotal swelling (hydrocoele). In 2000, WHO launched the Global Programme to Eliminate Lymphatic Filariasis (GPELF) to interrupt transmission through MDA and provide care for affected individuals. Between 2018 and 2020, São Tomé and Príncipe implemented nationwide treatment campaigns using WHO-recommended drug combinations.

To evaluate the reduction in transmission, surveys were conducted among adults in 2022 and again from 2024 to 2025. Very few people tested positive for infection: 21 out of 14 132 adults (0.15%) in 2022 and seven out of 14 653 (0.05%) in the 2024–2025 follow-up survey. No microfilariae were detected among antigen-positive individuals, and national records reported relatively low numbers of lymphoedema (190 cases) and hydrocoele (nine cases). Taken together, the findings indicate that ongoing transmission is very unlikely. They also suggest that parasite transmission is unlikely to persist and that São Tomé and Príncipe is close to eliminating lymphatic filariasis as a public health problem. However, it remains necessary to enhance services for the management of chronic disease manifestations.

## Introduction

Lymphatic filariasis (LF) is a mosquito-borne neglected tropical disease primarily caused by the filarial nematode *Wuchereria bancrofti* [1]. Chronic infection can lead to lymphoedema of the limbs and hydrocoele in adult males, both of which cause long-term disability, social stigma and economic hardship within affected communities [2]. Lymphatic Filariasis has historically affected hundreds of millions of people living in tropical and subtropical regions [3]. To address this burden, WHO established the GPELF in 2000 with the aim of eliminating the disease as a public health problem by interrupting transmission and providing morbidity management and disability prevention (MMDP) services for affected individuals [4–6].

The main strategy for interrupting transmission is annual MDA targeting at-risk populations [7]. Traditionally, MDA involved a two-drug regimen of albendazole combined with either ivermectin or diethylcarbamazine, depending on the epidemiological context. WHO has recently recommended the triple-drug regimen consisting of ivermectin, diethylcarbamazine, and albendazole (IDA) for use in settings where onchocerciasis and loiasis are absent [8]. Clinical and field studies have demonstrated that the IDA regimen eliminates microfilariae more rapidly than two-drug regimens and can accelerate progress towards elimination when high treatment coverage is achieved [9–13].

Monitoring and evaluation are essential components of the global elimination strategy because they provide evidence to inform programmatic decision-making and document progress towards elimination [4,6]. WHO recommends a series of epidemiological assessments following MDA to determine whether infection levels have declined to thresholds below which transmission is unlikely to persist [14]. In settings where the IDA regimen has been implemented, impact assessment surveys can provide valuable evidence on reductions in infection prevalence after treatment campaigns [14, 15].

São Tomé and Príncipe, an island nation in the Gulf of Guinea with a relatively small and geographically contained population, has historically reported evidence of LF transmission caused by *Wuchereria bancrofti*. Baseline mapping surveys conducted in 2015 using immunochromatographic antigen tests (ICT) identified positive cases across multiple districts and communities on both São Tomé and Príncipe islands, with between one and seven antigen-positive individuals detected per district, confirming endemic transmission prior to the initiation of MDA (ESPEN source: https://espen.afro.who.int/). In line with global elimination strategies, the Ministry of Health (MoH) implemented three consecutive rounds of nationwide MDA between 2018 and 2020 [16–18]. The first round in 2018 used the two-drug regimen of diethylcarbamazine and albendazole, followed by two rounds of the triple-drug (IDA) regimen in 2019 and 2020. The interventions were designed to quickly reduce the parasite reservoir in the human population and interrupt transmission.

To evaluate the impact of these interventions and guide programmatic decision-making, an IDA impact assessment survey was conducted in 2022 among adults aged 20 years and older using CFA detection. A follow-up survey was subsequently conducted between December 2024 and January 2025 to assess whether infection indicators remained below thresholds consistent with sustained transmission following cessation of MDA. This study describes the epidemiological findings from these assessments and documents the progress made by São Tomé and Príncipe towards validation of LF elimination as a public-health problem.

## Methodology

### Study setting

São Tomé and Príncipe is a small island nation in the Gulf of Guinea off the west coast of Central Africa. It consists of two main islands, São Tomé and Príncipe, with an estimated population of 220 000 inhabitants. Administratively, the country is divided into seven health districts: Água-Grande, Mé-Zóchi, Lobata, Cantagalo, Caué, Lembá, and the Autonomous Region of Príncipe. Its tropical climate, characterized by high humidity and rainfall, provides favourable ecological conditions for mosquito vectors such as *Anopheles* species, which are known to drive the transmission of *W. bancrofti* in São Tomé and Príncipe [18].

### National lymphatic filariasis elimination programme and MDA

After evidence of LF transmission was found in the country [19], (Fig. 1), the MoH implemented a national elimination programme aligned with the WHO GPELF strategy *(18).* MDA was conducted nationwide for three consecutive years. The first round in 2018 used a two-drug regimen consisting of diethylcarbamazine and albendazole. Subsequent rounds in 2019 and 2020 used the IDA triple-drug regimen in line with WHO recommendations aimed at accelerating interruption of transmission [20]. Treatment was delivered through community-based distribution targeting eligible populations across all health districts in the country.

**Fig. 1.**
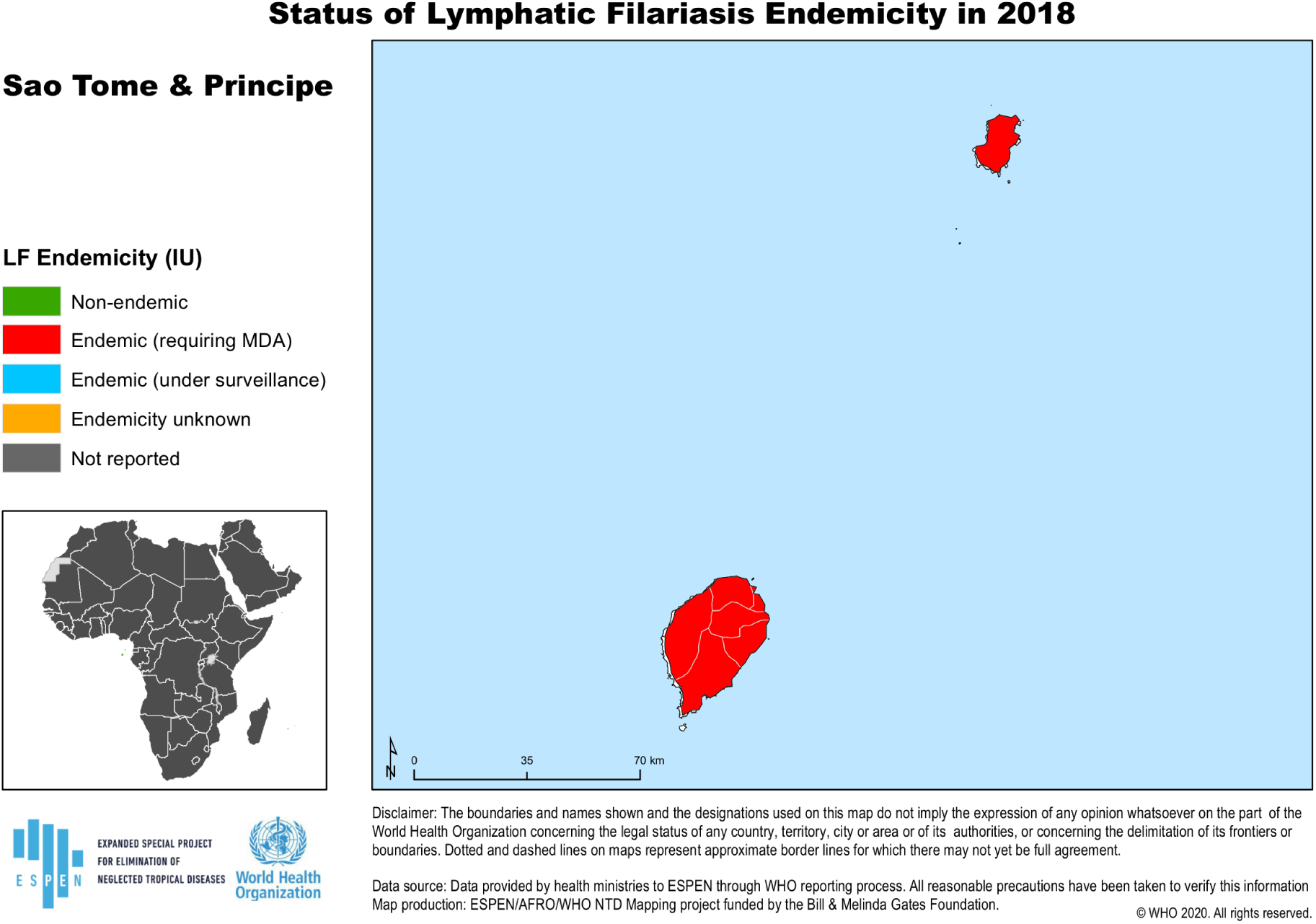
Pre-MDA endemicity status of lymphatic filariasis in São Tomé and Príncipe at implementation unit level (2018) (source: https://espen.afro.who.int/).

### IDA impact assessment surveys (2022 and 2024–2025)

To evaluate the impact of MDA and assess the persistence of infection following cessation of treatment, cross-sectional IDA impact assessment surveys were conducted between April and May 2022 and again between December 2024 and January 2025. Both surveys targeted adults aged 20 years and older, in line with WHO guidance for evaluating the impact of the IDA regimen [21].

The country was divided into five evaluation units by grouping districts with similar ecological, epidemiological, and population characteristics. Within each evaluation unit, communities were selected and households were visited to recruit eligible participants. CFA was detected through FTS testing of capillary blood obtained by finger prick. The FTS is a rapid diagnostic test widely used in LF surveillance programmes to detect circulating *W. bancrofti* antigen [22].

### Detection of microfilaremia

Individuals who tested positive for CFA during the surveys were invited to undergo nocturnal blood examination to detect microfilariae. Blood samples were collected at night, when *W. bancrofti* microfilariae are typically present in peripheral blood [14]. A calibrated thick blood smear was prepared from capillary blood, then examined under a microscope to detect and quantify microfilariae.

### Morbidity assessment

During community visits, individuals were also screened for clinical symptoms consistent with LF, including lymphoedema of the limbs and hydrocoele in adult males. Information on identified cases was recorded to document the burden of morbidity and support MMDP activities within the national programme.

### Data sources

The data used in this study were obtained from multiple sources within the MoH of São Tomé and Príncipe. MDA coverage data for 2018, 2019 and 2020 were extracted from the Joint Reporting Forms submitted annually by the MoH to WHO as part of routine programme monitoring (ESPEN source: https://espen.afro.who.int/). Data from the IDA impact assessment surveys conducted in 2022 and between December 2024 and January 2025 were obtained from official survey reports prepared by the MoH with technical support from the WHO Country Office and the Expanded Special Project for Elimination of Neglected Tropical Diseases (ESPEN). Data on morbidity cases consistent with LF, including lymphoedema and hydrocoele, were obtained from programme registers compiled during national programme activities.

### Data management and analysis

Survey data were recorded using standardized electronic data collection forms and entered into electronic databases for analysis. Descriptive statistics were used to summarize the number of individuals tested, CFA prevalence, the presence of microfilaremia among antigen-positive individuals and the distribution of morbidity cases across health districts.

### Ethics statement

The surveys described in this study were conducted as part of national LF elimination programme activities implemented by the MoH of São Tomé and Príncipe in collaboration with WHO. These activities formed part of routine public health surveillance used to monitor programme performance and guide elimination strategies [21]. Participation in the surveys was voluntary and individuals provided informed consent prior to testing. Personal identifiers were not included in the analytical datasets used for this study.

## Results

### MDA coverage (2018–2020)

Three consecutive rounds of MDA were implemented nationwide between 2018 and 2020. The first round in 2018 used the two-drug regimen of diethylcarbamazine and albendazole, while the second and third rounds in 2019 and 2020 used the IDA triple-drug regimen. Treatment was delivered in all seven health districts of São Tomé and Príncipe.

Epidemiological coverage exceeded the WHO-recommended threshold of 65% in all health districts during each round of treatment [23]. National coverage reached 86.4% in 2018, 74.2% in 2019, and 80.0% in 2020. Coverage varied slightly across health districts but remained consistently high throughout the programme period (Table 1).

**Table 1.** MDA coverage by district, São Tomé and Príncipe, 2018–2020.

| District | 2018 (DA) |  |  | 2019 (IDA) |  |  | 2020 (IDA) |  |  |
| --- | --- | --- | --- | --- | --- | --- | --- | --- | --- |
|  | Target | Treated | Coverage (%) | Target | Treated | Coverage (%) | Target | Treated | Coverage (%) |
| Água-Grande | 72 461 | 66 338 | 91.6 | 76 210 | 51 855 | 68 | 72 955 | 56 292 | 77.2 |
| Mé-Zóchi | 47 401 | 33 911 | 71.5 | 48 440 | 34 801 | 71.8 | 48 416 | 39 100 | 80.8 |
| Lobata | 20 508 | 18 635 | 90.9 | 21 015 | 16 204 | 77.1 | 21 075 | 16 059 | 76.2 |
| Cantagalo | 18 089 | 14 791 | 81.8 | 18 636 | 14 766 | 79.2 | 18 057 | 15 101 | 83.6 |
| Caué | 6 717 | 6 713 | 99.9 | 6 376 | 6 327 | 99.2 | 6 702 | 6 018 | 89.8 |
| Lembá | 15 177 | 14 192 | 93.5 | 16 099 | 12 357 | 76.8 | 14 929 | 12 288 | 82.3 |
| Príncipe | 7 851 | 7 931 | 101 | 7 894 | 8 217 | 104.1 | 7 938 | 7 261 | 91.5 |
| Total | 188 204 | 162 511 | 86.4 | 194 670 | 144 527 | 74.2 | 190 072 | 152 119 | 80 |

**DA:** Diethylcarbamazine and Albendazole; **IDA:** Ivermectin, Diethylcarbamazine and Albendazole

### IDA impact assessment survey (2022)

The first IDA impact assessment survey, conducted in 2022, examined 14 132 adults aged 20 years and older across five evaluation units. CFA was detected in 21 individuals, corresponding to an overall antigen positivity of 0.15%.

Antigen-positive individuals were identified in all evaluation units. The highest number of positive individuals was detected in the Caué–Lembá evaluation unit (eight cases), followed by Lobata–Mé-Zóchi (seven cases). Príncipe and Cantagalo reported three and two positive individuals respectively, while Água-Grande reported one positive individual.

In all the evaluation units, the number of antigen-positive individuals remained below the critical cut-off values defined in the WHO decision framework [14] thresholds designed to ensure that infection prevalence is sufficiently low (generally around 1–2%) to prevent ongoing transmission – indicating that transmission is unlikely to be sustained (Table 2).

**Table 2.** Results of IDA impact assessment surveys in adults aged 20 years and older, São Tomé and Príncipe, 2022 and 2024–2025.

| Evaluation Unit | Health Districts Included | Survey 2022 |  | Survey 2024–2025 |  | Critical Cut-off | Interpretation |
| --- | --- | --- | --- | --- | --- | --- | --- |
|  |  | Adults examined | Antigen positive | Adults examined | Antigen positive |  |  |
| Príncipe | Príncipe | 1 509 | 3 | 1 589 | 3 | 9 | Below threshold |
| Cantagalo | Cantagalo | 3 116 | 2 | 3 581 | 2 | 18 | Below threshold |
| Caué–Lembá | Caué, Lembá | 3 178 | 8 | 3 282 | 1 | 18 | Below threshold |
| Lobata–Mé-Zóchi | Lobata, Mé-Zóchi | 3 203 | 7 | 3 006 | 1 | 18 | Below threshold |
| Água-Grande | Água-Grande | 3 126 | 1 | 3 195 | 0 | 18 | Below threshold |
| **Total** | — | **14 132** | **21** | **14 653** | **7** | — | Transmission unlikely |

### Follow-up impact assessment survey (2024–2025)

A follow-up survey conducted between December 2024 and January 2025 examined 14 653 adults aged 20 years and older across the same evaluation units. Seven individuals tested positive for CFA, corresponding to an overall antigen positivity of approximately 0.05%. Antigen-positive individuals were detected in four evaluation units: Principe (three cases), Cantagalo (two cases), Caué–Lembá (one case), and Lobata–Mé-Zóchi (one case). No positive individuals were detected in Água-Grande. All evaluation units remained below the WHO decision thresholds, confirming the findings from the initial IDA impact assessment survey and reinforcing the conclusion that LF transmission is unlikely to persist in the population (Table 2).

### Microfilaremia findings

All individuals who tested positive for CFA during the two surveys were invited to undergo nocturnal calibrated thick blood smear microscopy to detect microfilariae. A total of 28 antigen-positive individuals were examined, including 21 identified in 2022 and seven identified during the 2024–2025 survey. No microfilaria was detected in any of the blood smears examined, indicating the absence of detectable *W. bancrofti* microfilaremia among the antigen-positive individuals.

### Morbidity findings

Clinical screening and programme data identified individuals with chronic symptoms of LF in all health districts of São Tomé and Príncipe. During the 2022 IDA impact assessment survey, 106 individuals with lymphoedema and 9 with hydrocoele were identified. An updated national morbidity register compiled in 2024 recorded an increase to 190 lymphoedema cases, while the number of hydrocoele cases remained unchanged at nine (Table 3).

**Table 3.** Distribution of lymphoedema and hydrocoele cases identified during the 2022 IDA impact assessment survey and recorded in the national morbidity register (2024), São Tomé and Príncipe.

| District | Lymphoedema |  | Hydrocoele |  |
| --- | --- | --- | --- | --- |
|  | 2022 | 2024 | 2022 | 2024 |
| Água-Grande | 8 | 48 | 1 | 1 |
| Mé-Zóchi | 9 | 52 | 0 | 2 |
| Lobata | 10 | 28 | 1 | 1 |
| Cantagalo | 15 | 17 | 0 | 1 |
| Caué | 27 | 12 | 0 | 2 |
| Lembá | 26 | 21 | 1 | 1 |
| Príncipe | 11 | 12 | 6 | 1 |
| **Total** | **106** | **190** | **9** | **9** |

Lymphoedema cases were reported in all health districts in both datasets. In the 2022 survey, higher concentrations were found in Caué and Lembá, while the 2024 register showed more cases in Mé-Zóchi and Água-Grande. Hydrocoele cases were less common and more geographically concentrated, with a notable cluster in Príncipe during the 2022 survey and a more even distribution across health districts in 2024 (Table 3).

## Discussion

The GPELF is built on two mutually reinforcing pillars: interrupting transmission through preventive chemotherapy and alleviating suffering via MMDP [7, 24, 25]. The findings of this study indicate that São Tomé and Príncipe has made substantial progress towards achieving the first pillar. Three consecutive rounds of nationwide MDA implemented between 2018 and 2020 consistently achieved high epidemiological coverage across all health districts, exceeding the thresholds required to interrupt transmission [18]. The subsequent introduction of the IDA regimen in 2019 and 2020 likely helped accelerate parasite elimination and reduce infection levels faster than the traditional two-drug regimens [12, 13].

The results of the IDA impact assessment surveys conducted in 2022 and again between December 2024 and January 2025 showed extremely low levels of CFA across all evaluation units. In addition, nocturnal calibrated thick blood smear microscopy performed on antigen-positive individuals did not detect *W. bancrofti* microfilariae. The findings suggest that parasite transmission has declined to levels unlikely to sustain the parasite life cycle in the population. Similar declines in infection indicators have also been reported in other endemic settings following the introduction of the IDA regimen which aims to accelerate progress towards elimination [10, 26–28].

Several factors may have contributed to the progress observed in São Tomé and Príncipe. Experience from other elimination programmes has shown that sustained political commitment, strong programme leadership and high treatment coverage are key determinants of successful LF elimination efforts [29–32]. In São Tomé and Príncipe, the nationwide implementation of MDA, engagement of community health workers, and close collaboration between the MoH and international partners such as WHO facilitated effective programme implementation. The relatively small population size and insular geographic setting– similar to small island settings in the Western Pacific such as Niue, the Cook Islands and Samoa – may also have contributed to the rapid reduction of infection by limiting population movement and facilitating high nationwide treatment coverage [31, 33, 34].

Vector control interventions implemented as part of the national malaria elimination programme may also have contributed to the observed reduction in LF transmission. As the country strives to eliminate malaria by 2025, integrated strategies including vector control, early diagnosis, MDA with artemisinin-piperaquine plus primaquine in residual hotspots and effective case management have been scaled up [35–38]. Measures targeting *Anopheles* mosquitoes – such as larval source management using *Bacillus thuringiensis israelensis* (Bti), indoor residual spraying (IRS) and widespread use of long-lasting insecticidal nets (LLINs) – have achieved substantial coverage [39, 40]. These interventions likely reduced human–vector contact and may have had a collateral impact on LF transmission.

While the programme has made significant progress towards interrupting transmission, the implementation of the second pillar of the elimination strategy, MMDP, remains limited. Individuals with chronic symptoms of LF, such as lymphoedema and hydrocoele, have been identified during programme activities. However, structured and accessible services for these individuals are not yet systematically integrated into the national health system. Addressing morbidity is a core requirement for validation of elimination, as the global strategy aims not only to interrupt transmission but also to alleviate suffering among affected individuals [41–43]. The national programme should therefore prioritize establishing and expanding morbidity management services across all health districts, including standardized care for lymphoedema and access to hydrocoele surgery. In parallel, documenting the availability, accessibility and quality of these services through facility assessments will be essential to demonstrate that MMDP components are functional and meet validation requirements.

Progress in this area has also been hampered by operational disruptions. Morbidity management activities initiated prior to the COVID-19 pandemic – particularly in Lobata and Lembá, where health personnel were trained and self-care materials were distributed – were disrupted and have not yet been fully reinstated nationwide. Strengthening efforts to resume case identification and expand service delivery will therefore be critical, both to improve patient care and to ensure that the programme can generate the evidence required for the national elimination dossier.

The timing of surveillance activities in São Tomé and Príncipe should also be aligned with WHO guidance, which recommends an interval of approximately four years between cessation of MDA and the final assessment of transmission [14]. Since MDA ended in 2020, the timing for this assessment was expected to be around 2024. Although there was a slight delay in implementing the IDA impact assessment surveys, with the latest survey being conducted between December 2024 and January 2025, the findings consistently indicate very low infection levels, supporting the conclusion that transmission has likely been interrupted.

Taken together, evidence from routine programme monitoring and the two IDA impact assessment surveys indicates that São Tomé and Príncipe is nearing the threshold required for validation of elimination of LF as a public health problem. The programme should now prioritize strengthening MMDP services while advancing the preparation of the national elimination dossier.

The findings indicate that São Tomé and Príncipe has made substantial progress towards eliminating LF as a public health problem. High coverage levels achieved during three rounds of nationwide MDA between 2018 and 2020, followed by consistently low CFA levels and no detectable microfilaremia in post-MDA impact assessments conducted in 2022 and 2024–2025, suggest that transmission has likely been reduced to levels insufficient to sustain the parasite lifecycle. As the programme progresses towards validation, priority should now be given to strengthening MMDP services, including access to lymphoedema care and hydrocoele surgery, while also documenting service availability and quality. Advancing these efforts, together with preparing the national elimination dossier, will be essential to support formal validation by WHO.

## Acknowledgements

The authors acknowledge the Ministry of Health of São Tomé and Príncipe for its leadership and coordination of LF activities. We thank the national and district health teams who conducted the MDA campaigns and the IDA impact assessment surveys. We also acknowledge the technical support provided by the WHO Regional Office for Africa through the WHO Country Office and ESPEN, as well as the contributions of the field teams, laboratory personnel and community health workers who helped implement the surveys.

## Authors’ contributions

Conceptualization: DB, JK, EJ, AR.

Methodology: DB, JC.

Investigation: ASV, JPK, JPT, AR.

Data curation: DY, HMZ.

Formal analysis: DB, JC.

Writing – original draft: DB, JC.

Writing – review and editing: DB, JC, ASV, JPK, JPT, DY, HMZ, JK, EJ,AR,JVF,AD,LD,VSG

## Data availability

All relevant data supporting the findings of this study are included within the manuscript.

## Competing Interests

The authors have declared that no competing interests exist.

## Notes

### Competing Interest Statement

The authors have declared no competing interest.

### Funding Statement

The author(s) received no specific funding for this work.

### Author Declarations

The NTD program in Sao Tome e Principle obtained approval from the Government to conduct surveys. WHO staff obtained a clearance (Executive clearance granted to Product ID -AF-2026-01393 to publish this study. Participation in the surveys was voluntary and individuals provided informed consent prior to testing. Personal identifiers were not included in the analytical datasets used for this study.

